# Variants in *ACTC1* underlie distal arthrogryposis accompanied by congenital heart defects

**DOI:** 10.1101/2023.03.07.23286862

**Authors:** Jessica X. Chong, Matthew Carter Childers, Colby T. Marvin, Anthony J. Marcello, Hernan Gonorazky, Lili-Naz Hazrati, James J. Dowling, Fatema Al Amrani, Yasemin Alanay, Yolanda Nieto, Miguel Á Marín Gabriel, Arthur S. Aylsworth, Kati J. Buckingham, Kathryn M. Shively, Olivia Sommers, Kailyn Anderson, University of Washington Center for Mendelian Genomics, University of Washington Center for Rare Disease Research, Michael Regnier, Michael J. Bamshad

## Abstract

Contraction of the human sarcomere is the result of interactions between myosin cross-bridges and actin filaments. Pathogenic variants in genes such as *MYH7*, *TPM1*, and *TNNI3* that encode parts of the cardiac sarcomere cause muscle diseases that affect the heart, such as dilated cardiomyopathy and hypertrophic cardiomyopathy. In contrast, pathogenic variants in homologous genes *MYH2*, *TPM2*, and *TNNI2*, that encode parts of the skeletal muscle sarcomere, cause muscle diseases affecting skeletal muscle, such as the distal arthrogryposis (DA) syndromes and skeletal myopathies. To date, there have been few reports of genes (e.g., *MYH7*) encoding sarcomeric proteins in which the same pathogenic variant affects both skeletal and cardiac muscle. Moreover, none of the known genes underlying DA have been found to contain mutations that also cause cardiac abnormalities. We report five families with DA due to heterozygous missense variants in the gene *actin, alpha, cardiac muscle 1* (*ACTC1*). *ACTC1* encodes a highly conserved actin that binds to myosin in both cardiac and skeletal muscle.

Mutations in *ACTC1* have previously been found to underlie atrial septal defect, dilated cardiomyopathy, hypertrophic cardiomyopathy, and left ventricular noncompaction. Our discovery delineates a new DA condition due to mutations in *ACTC1* and suggests that some functions of *actin, alpha, cardiac muscle 1* are shared in cardiac and skeletal muscle.

## Introduction

Sarcomeres are the repeating functional units of muscle cells that are joined end-to-end to form skeletal and cardiac muscle fibers.^1^ Sarcomeres consist of thick myosin filaments and thin actin filaments along with proteins such as troponin and tropomyosin that facilitate and regulate the interactions between the filaments.^2^ Contractile force is generated when the myosin and actin filaments bind to form cross-bridges, which is thought to cause the filaments to slide against each other. The key sarcomeric proteins are encoded by highly conserved and homologous genes that typically express an isoform that is predominant in either cardiac or skeletal muscle. For example, *TNNI2, MYH3*, and *ACTA1* encode isoforms of troponin, myosin heavy chain, and alpha actin, respectively, that are primarily expressed in skeletal muscle while *TNNI3*, *MYH6*, and *ACTC1* encode isoforms that are primarily expressed in cardiac muscle.^3^

Pathogenic variants in genes that encode the skeletal sarcomeric proteins tropomyosin (*TPM2* [MIM: 190990]);^4^ troponin I2, fast skeletal type (*TNNI2* [MIM: 191043]);^4^ troponin T3, fast skeletal type (*TNNT3* [MIM: 600692]),^5^ myosin heavy chain 3 (*MYH3* [MIM:160720]),^6^ and myosin heavy chain 8 (*MYH8* [MIM: 160741])^7^ account for most cases of distal arthrogryposis (DA), a group of Mendelian conditions characterized by non-progressive congenital contractures of the limbs, and less frequently, contractures of the face, ocular muscles, neck webbing, pterygia, short stature, and scoliosis. The precise pathogenesis of the contractures is unknown, although it has been proposed that pathogenic variants lead to perturbation of muscle contraction or relaxation, resulting in reduced limb movement in utero.^8, 9^ Several additional genes, specifically *PIEZO2* [MIM: 613629],^10, 11^ *ECEL1* [MIM: 605896],^12, 13^ and *FBN2* [MIM:612570],^14^ that underlie other forms of DA encode proteins that are less directly involved in sarcomere contraction.

To date, the vast majority of variants in genes encoding homologous components of the cardiac sarcomere have been found to result in conditions in which only cardiac muscle is affected, including cardiomyopathy and structural heart defects. Herein, we report five families with DA and congenital heart defects due to heterozygous missense variants in the gene *actin, alpha, cardiac muscle 1* (*ACTC1*)*. ACTC1* encodes a highly conserved actin that binds to myosin in both cardiac and skeletal muscle. We employ molecular dynamics (MD) simulations of wild type (WT) and mutant cardiac actin to predict the structural and functional consequences of these variants.

## Methods

### Exome Sequencing, Annotation, and Filtering

From a cohort of 463 families (1,582 individuals) with multiple congenital contractures, we selected 172 families, in which pathogenic or likely pathogenic variants had not been identified, for exome sequencing (ES). All studies were approved by the institutional review boards of the University of Washington and Seattle Children’s Hospital and informed consent was obtained from each participant or their parents to participate in this study and this publication. ES, annotation, and analysis were performed by the University of Washington Center for Mendelian Genomics (now the University of Washington Center for Rare Disease Research) as described previously.^15^ Briefly, variants were called by GATK v3.7 HaplotypeCaller and annotated with Variant Effect Predictor v95.3.^16^ Variants were filtered using GEMINI v0.30.1^17^ for genotype call quality (GQ≥20), read depth (≥6), allele frequency in population controls (i.e., maximum frequency in any continental superpopulation in gnomAD^18^ v2.1 and v3.0 exomes and genomes <0.005), consistency with the mode of inheritance in each family, and predicted impact on protein-coding sequence (e.g. annotated as missense, nonsense, canonical splice, or coding indel).

### Molecular Dynamics – Model Preparation and Simulation

Initial coordinates for cardiac globular (g-actin) structures were generated via homology to an X-ray crystal structure of rabbit skeletal actin (UniProt Entry P68135) downloaded from the Protein Data Bank^19^ (PDB, www.rcsb.org ID 3HBT)^20^. 3HBT is a model of g-actin complexed with ATP, CA, and SO4. The human ACTC1 sequence was downloaded from UniProt (Entry P68032). The human and rabbit sequences were 98.9 % identical, assessed using *Clustal Omega*^21^ and there were four conservative amino acid substitutions ([human amino acid, human residue number, rabbit amino acid as follows]: D2E, E3D, L301M, and S360T). Homology models of the human wild type (WT) and four mutant (p.Thr68Asn, p.Arg185Trp, p.Gly199Ser, p.Arg374Ser) structures were generated using *Modeller*^22^, which introduced the amino acid substitutions and built coordinates for atoms not present in the PDB file (no coordinates were present for D-loop residues 40-50 in 3HBT). The p.Arg374His variant was not simulated due to the expected overlap with p.Arg374Ser and because the change to His is more conservative than the change to Ser. For p.Gly199Ser, the initial backbone dihedral (φ,ψ) angles for Gly were (152°, -16°), which are unfavorable for Ser. Consequently, this loop was further refined and the initial S199 dihedrals were (-177°, -14°). During modelling, crystallographic waters and the SO4 were removed, ATP was retained, and Ca2+ was replaced by Mg^2+^. Initial coordinates for cardiac filamentous (f-actin) pentamer structures were generated using an electron microscopy structure of mouse tropomyosin and rabbit skeletal actin (PDB ID: 3J8A).^23^ The tropomyosin chains were removed and the f-actin pentamers complexed with ADP and Mg^2+^ were used to construct homology models of WT and p.Thr68Asn human cardiac F-actin with *Modeller.* After homology models were built, hydrogen atoms were modeled onto the initial structure using the *tleap* module of AMBER and each protein was solvated with explicit water molecules in a periodic, truncated octahedral box that extended 10 Å beyond any protein atom. Na^+^ counterions were added to neutralize the systems.

All simulations were performed with the AMBER20 package^24, 25^ and the ff14SB force field^26^ using standard procedures. Water molecules were treated with the TIP3P force field.^27^ Metal ions were modeled using the Li and Merz parameter set.^28–30^ ATP and ADP molecules were treated with parameters from Meagher et al.^31^ The SHAKE algorithm was used to constrain the motion of hydrogen-containing bonds. Long-range electrostatic interactions were calculated using the particle mesh Ewald (PME) method. Each system was minimized in for 10000 steps divided across three stages in which restraints were placed either on hydrogen atoms, solvent atoms, or all backbone heavy atoms (C_α_, C, N, O atoms). After minimization, systems were heated to 310 K over 300 ps using the canonical NVT (constant number of particles, volume, and temperature) ensemble. During all heating stages, 25 kcal mol-1 restraints were present on the backbone heavy atoms (C_α_, C, N, O atoms). After the system temperatures reached 310 K, the systems were equilibrated for 5.4 ns over 5 successive stages using the NPT (constant number of particles, pressure, and temperature) ensemble. During equilibration, restraints on backbone atoms were decreased from 25 kcal mol^-1^ during the first stage to 1 kcal mol^-1^ during the fourth stage. During the final equilibration stage, the systems were equilibrated in the absence of restraints. Production dynamics for conventional molecular dynamics (cMD) simulations were then performed using the NVT ensemble using an 8 Å nonbonded cutoff, a 2 fs time step, and coordinates were saved every picosecond. G-actin cMD simulations were run in triplicate each replicate simulation was 500 ns long. We used an enhanced sampling scheme called Gaussian accelerated MD (GaMD)^32^ to explore conformational sampling in the WT and p.Thr68Asn F-actin pentamer models. GaMD production runs were preceded by a 52 ns long GaMD equilibration period in which boost potentials were added. The upper limits of the standard deviation of the boost potentials were set to 6 kcal mol^-1^. Neither the standard 5.2 ns equilibration nor the 52 ns GaMD equilibration contributed to the length of the production dynamics for any simulation. Production dynamics for Gaussian accelerated (GaMD) simulations were using the NVT ensemble using an 8 Å nonbonded cutoff, a 2 fs time step, and coordinates were saved every picosecond. Single replicates of the WT and p.Thr68Asn were performed and each simulation was 300 ns long. Unless specified otherwise, simulations were analyzed separately, and the results of replicate simulations were averaged together.

### Molecular Dynamics – Analysis

The C_α_ root-mean-squared deviation (RMSD), C_α_ root-mean-squared fluctuation (RMSF), solvent accessible surface area (SASA), secondary structure content, inter-atomic distances, and inter-residue contacts were calculated with *cpptraj*.^33^ The C RMSD was calculated after alignment of all C_α_ atoms to the minimized structure. The C_α_ RMSF was calculated about average MD structures for each simulation. For each timepoint in the simulation, two residues were considered in contact with one another if at least one pair of heavy atoms were within 5 Å of one another. Then we recorded the average percent simulation time each residue pair was in contact for each simulation. A student’s t-test was used to identify statistically significant (p < 0.05) differences in inter-residue contact times between the WT and mutant simulations. All protein images were prepared using *UCSF Chimera*^34, 35^.

## Results

After variant filtration of the exome data, three families had compelling candidate variants in the same candidate gene, *ACTC1,* or *actin, alpha, cardiac muscle* (*ACTC1* [MIM 102540; Refseq accession NM_005159.4]) (Table 1; Figure 1; Figure S1). Specifically, each family had a heterozygous candidate missense variant that was either *de novo* or segregated in an autosomal dominant pattern. In Family A, comprised of an affected father and affected daughter with camptodactyly of the fingers, hypoplastic flexion creases, clubfoot, webbed neck, scoliosis, hip contractures, and ventriculoseptal defect, a heterozygous variant in *ACTC1* (c.595G>A, p.Gly199Ser) was identified. This family was previously described (Family D in ^36^) as possibly having autosomal dominant multiple pterygium syndrome (MIM 178110), but no likely pathogenic or pathogenic variants in *MYH3* were identified. In Family B, an affected mother and affected daughter with knee contractures, clubfoot, limited neck rotation, scoliosis, and hip contractures, were heterozygous for c.1120C>A, p.Arg374Ser. The grandmother in Family B was described as having similar clinical findings but no medical records or photographs were available. Family C, the third family, was a simplex family in which a *de novo* variant c.203C>A, p.Thr68Asn was identified in the proband with clubfoot, camptodactyly of the toes and fingers, webbed neck, and an atrial septal defect. Upon follow up with Family C approximately twenty years after they were originally enrolled, the proband was found to have a son who had camptodactyly of the fingers, overlapping toes, elbow contractures, elbow webbing, and an atrial septal defect. Her son was heterozygous for the c.203C>A, p.Thr68Asn variant.

**Table 1.**
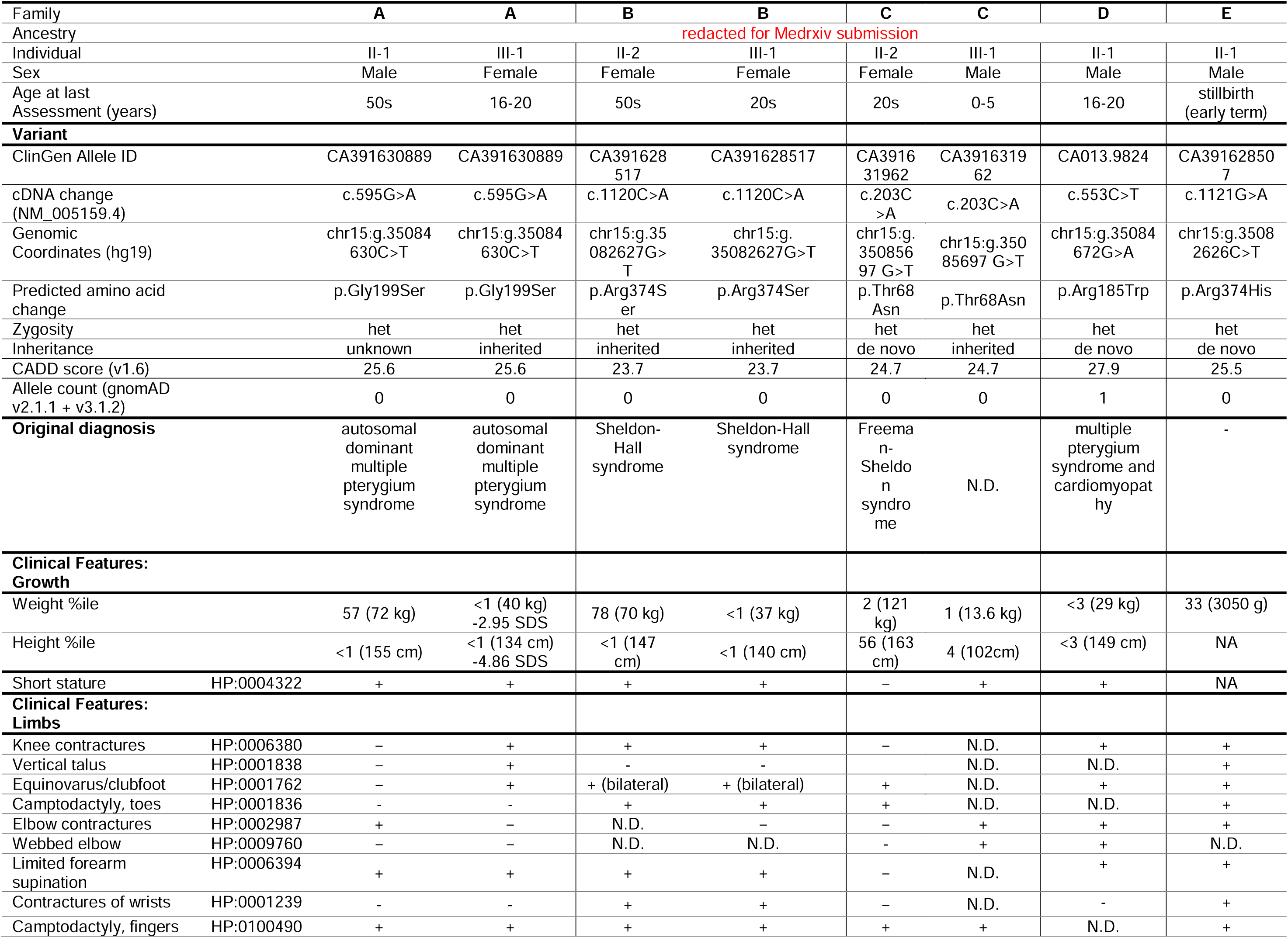
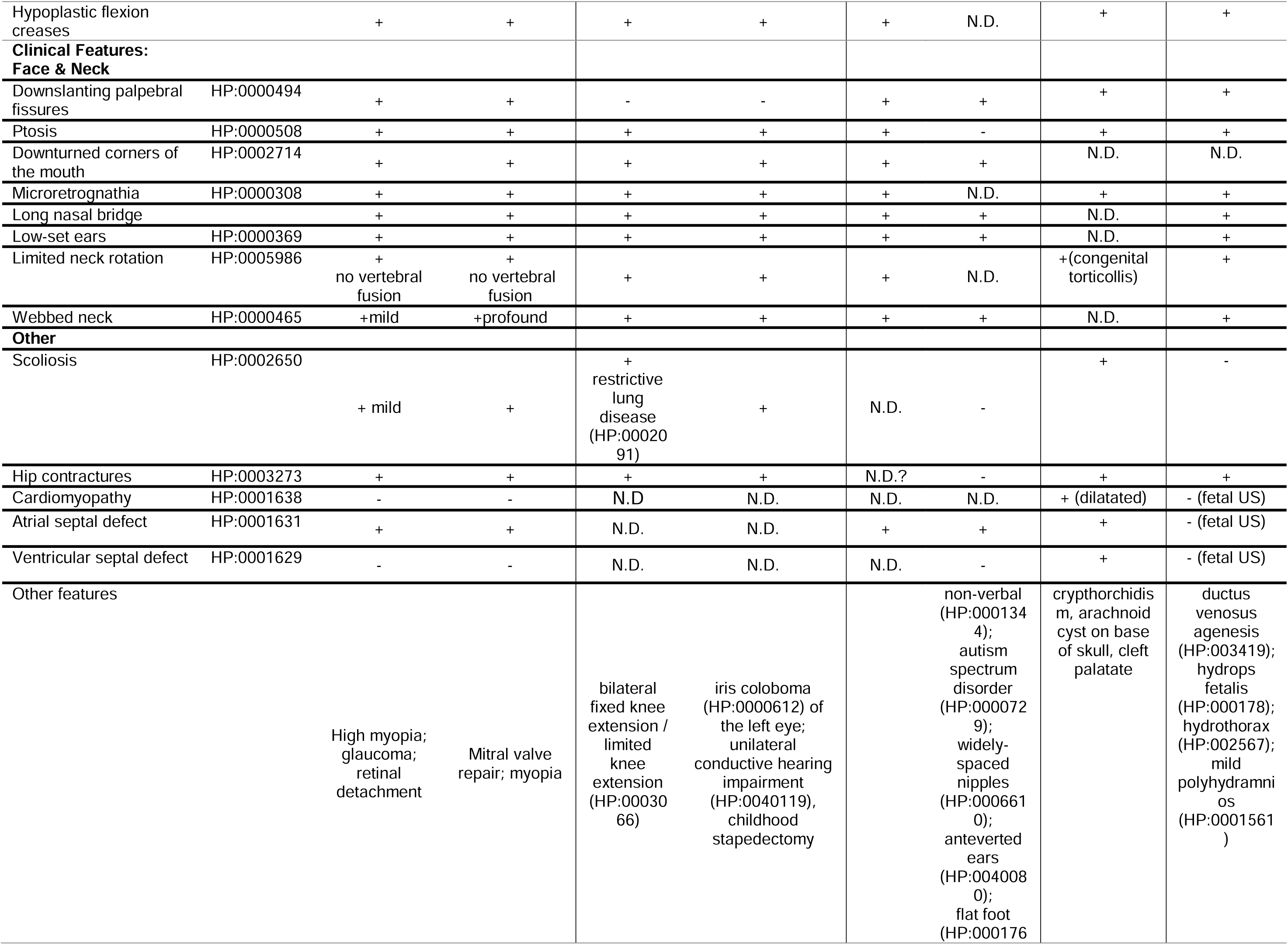
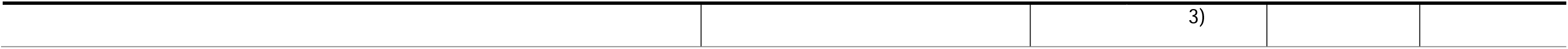
Clinical findings of individuals with Distal Arthrogryposis due to heterozygous variants in ACTC1. Plus (+) indicates presence of a finding, minus (-) indicates absence of a finding. * = described per report. ND = no data were available. NA = not applicable. CADD = Combined Annotation Dependent Depletion v1.6. cDNA positions named using HGVS notation and RefSeq transcript NM_005159.4. Predicted amino acid changes are shown. US = Ultrasound.

**Figure 1.**
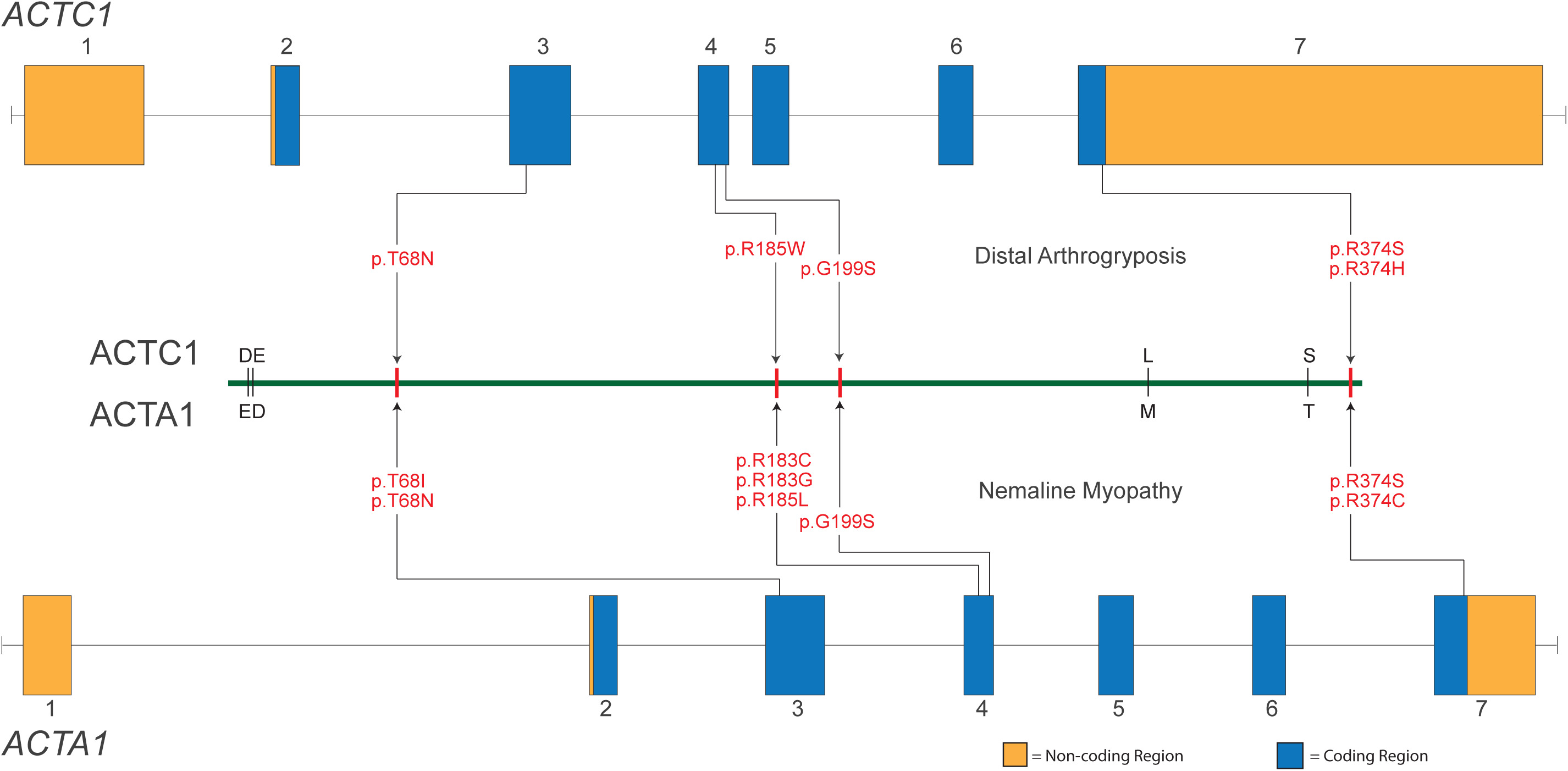
Phenotypic Characteristics of Individuals with Distal Arthrogryposis due to heterozygous variants in *ACTC1*. <u>Photo is redacted for Medrxiv submission.</u> Characteristics shown include: webbed neck, bilateral clubfoot, camptodactyly of the fingers and hypoplastic flexion creases in Family A (II-1 and III-1); camptodactyly, webbed neck, bilateral clubfoot, camptodactyly of the fingers and toes and hypoplastic flexion creases in Family B (II-2 and III-1); webbed neck, bilateral clubfoot, webbed neck, bilateral clubfoot, camptodactyly of the fingers and toes in Family C (II-2); and ptosis, webbed neck, camptodactyly of the fingers, and scoliosis in Family D (II-1). Table 1 contains a detailed description of the clinical findings of each affected individual and Figure S1 provides a pedigree for each family.

Two additional simplex families (Families D and E) subsequently came to our attention via direct clinician referral after clinical testing identified *ACTC1* as a candidate gene. Each had a *de novo* variant, c.553C>T, p.Arg185Trp in Family D, and c.1121G>A, p.Arg374His in Family E. Notably the variants in Family B and E perturbed the same residue. The proband of Family D had short stature, clubfoot, knee and elbow contractures, hypoplastic flexion creases, atrial septal defect, and ventricular septal defect. No muscle weakness was noted. An echocardiogram found borderline left ventricular systolic function with an ejection fraction of 52% and reduced longitudinal strain in the basal wall segments. Electron microscopy and immunohistochemistry of a skeletal muscle biopsy revealed possible nemaline bodies (Figure S2) and presence of rare myofibers with central core but no ragged red fibers or rod-like inclusions. The proband in Family E was stillborn at 38+3 weeks. At 31 weeks, ultrasound findings included hydrops fetalis, hydrothrorax, small lower jaw, ductus venosus agenesis, and positioning of the extremities consistent with fetal akinesia. No additional clinical information was available.

In summary, at least one affected individual in each family was reported to have a combination of camptodactyly of the fingers or toes, hypoplastic flexion creases, clubfoot, limited neck rotation, scoliosis (excluding Family E, for which only fetal ultrasound was available), and hip contractures. Common facial features included microretrognathia, ptosis, downslanting palpebral fissures, low-set ears, and a long nasal bridge (Figure 1). Ventricular or atrial septal defects were reported in families A, C, and D while E had ductus venosus agenesis in utero, but only one affected individual, the proband in D, had cardiomyopathy. The co-occurrence of these congenital heart defects is notable because *ACTC1* is well-established to underlie isolated cardiac abnormalities including dilated and hypertrophic cardiomyopathy (MIM 613424, 612098), atrial septal defects (MIM 612794), and left ventricular noncompaction (MIM 613424). However, *ACTC1* has not been reported to underlie a multiple malformation syndrome that affects multiple organ syndromes.

CADD (v1.6)^37^ scores >20.0 indicate that all five variants are predicted to be pathogenic (Table 1). For all five of these variants, the homologous residues in ACTA1 have been reported^38–43^ to be perturbed in infants with autosomal dominant severe congenital nemaline myopathy (Figure 2) leading to death before one year of age, an observation that further suggests these residues play a critical role in sarcomere function. In addition, these variants were either absent or exceedingly rare in gnomAD v2.1.1 or v3.1.2. p.Arg185Trp was heterozygous in a single individual in gnomAD and was the only variant that has been previously reported in ClinVar (twice classified as “likely pathogenic”). One of these ClinVar entries (SCV000742090.2) reports that p.Arg185Trp was found in an individual with “arthrogryposis multiplex congenita, multiple suture craniosynostosis, high palate, cleft uvula, pulmonary hypoplasia, bronchomalacia, pulmonary arterial hypertension, hydrocephalus, cryptorchidism, penile hypospadias, dysphagia, secundum atrial septal defect, patent foramen ovale, shallow orbits, infra-orbital crease, microretrognathia, webbed neck, short neck.” These clinical findings suggest that this individual likely has the same condition we describe herein.

**Figure 2.**
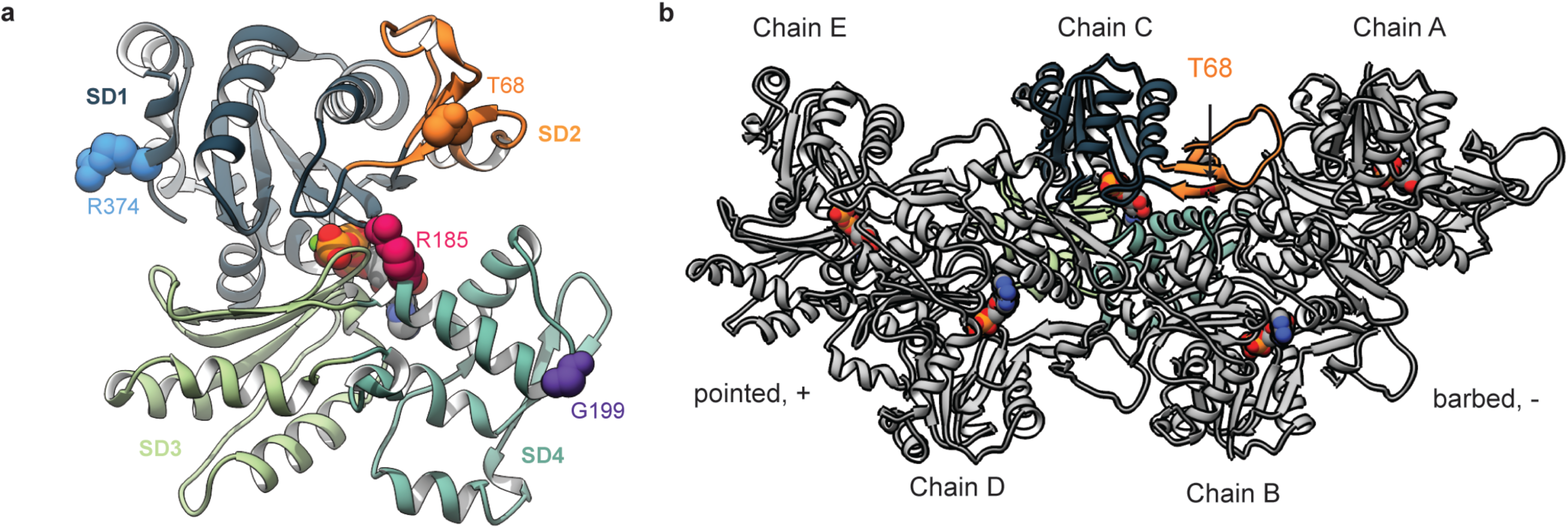
Genomic Model of *ACTC1* and *ACTA1*. Illustrated are each of the variants found in ACTC1 that underlie distal arthrogryposis and the homologous sites in ACTA1 that result in severe nemaline myopathy when mutated. *ACTC1* and *ACTA1* are each composed of 7 exons and consist of protein-coding (blue) and non-coding (orange) sequence. The proteins are nearly identical except for four residues (represented by single-letter amino acid codes immediately above and below the green line). The approximate location of each pathogenic variant (red text) is indicated by an arrow.

### Mutation-associated changes in the overall conformation and dynamics of g-actin

We first analyzed the conformations sampled by WT and mutant g-actin to determine whether four of the variants we identified (Figure 3A) led to large-scale conformational changes within actin monomers. We calculated the C_α_ root-mean-squared deviation (C_α_ RMSD, a measure of structural similarity) of each frame in the simulation to the minimized structure. The overall conformation of the actin monomers was preserved despite introduction of each mutation. In fact, the mutant simulations all had smaller C_α_ RMSD values than the WT simulations, indicating that the mutations dampened structural fluctuations in g-actin. All simulated systems had an average C_α_ root-mean-squared deviation (a measure of structural similarity) less than 2.6 Å to the crystallographic conformation (Table 2, WT: 2.6 Å, p.p.Thr68Asn: 2.0 Å test statistic=5.09, p=0.007, p.Gly199Ser, p.Arg185Trp: 2.4 Å test statistic=2.00, p=0.12, p.Gly199Ser: 2.4 Å test statistic=1.14, p=0.32, p.Arg374Ser: 2.2 Å test statistic=5.33, p =0.006; p-values denote statistical differences in the average RMSD values of the WT simulations versus each mutant).The largest amplitude structural change was a breathing motion in which relative scissoring of SD2 and SD4 opened and closed the nucleotide binding pocket, which occurred in all simulations. We next measured the C_α_ root-mean-squared fluctuations (C_α_ RMSF) and compared them with the WT simulations (Figure S3). The majority of residues in g-actin had small ( < 1 Å) C_α_ fluctuations about their average positions. The regions with the greatest fluctuations were the DNase1 binding loop (aka the D-loop, residues 41-56) and two loops in SD4 (residues 199-204, 219-224). The flexibilities of most residues were not affected by the mutations. However, all four mutations led to a decrease in the C_α_ RMSF of residues in the D-loop (Figure S3), and statistically significant (residues with significant differences denoted in Figure S3) decreases in the C_α_ RMSF of D-loop residues were observed for p.Thr68Asn, p.Arg185Trp, and p.Gly199Ser. Each of the mutations also caused low magnitude (< 0.5 Å) but statistically significant (residues with significant differences denoted in Figure S3) (p value and test) changes in C_α_ RMSF among residues near the mutation sites.

**Figure 3.**
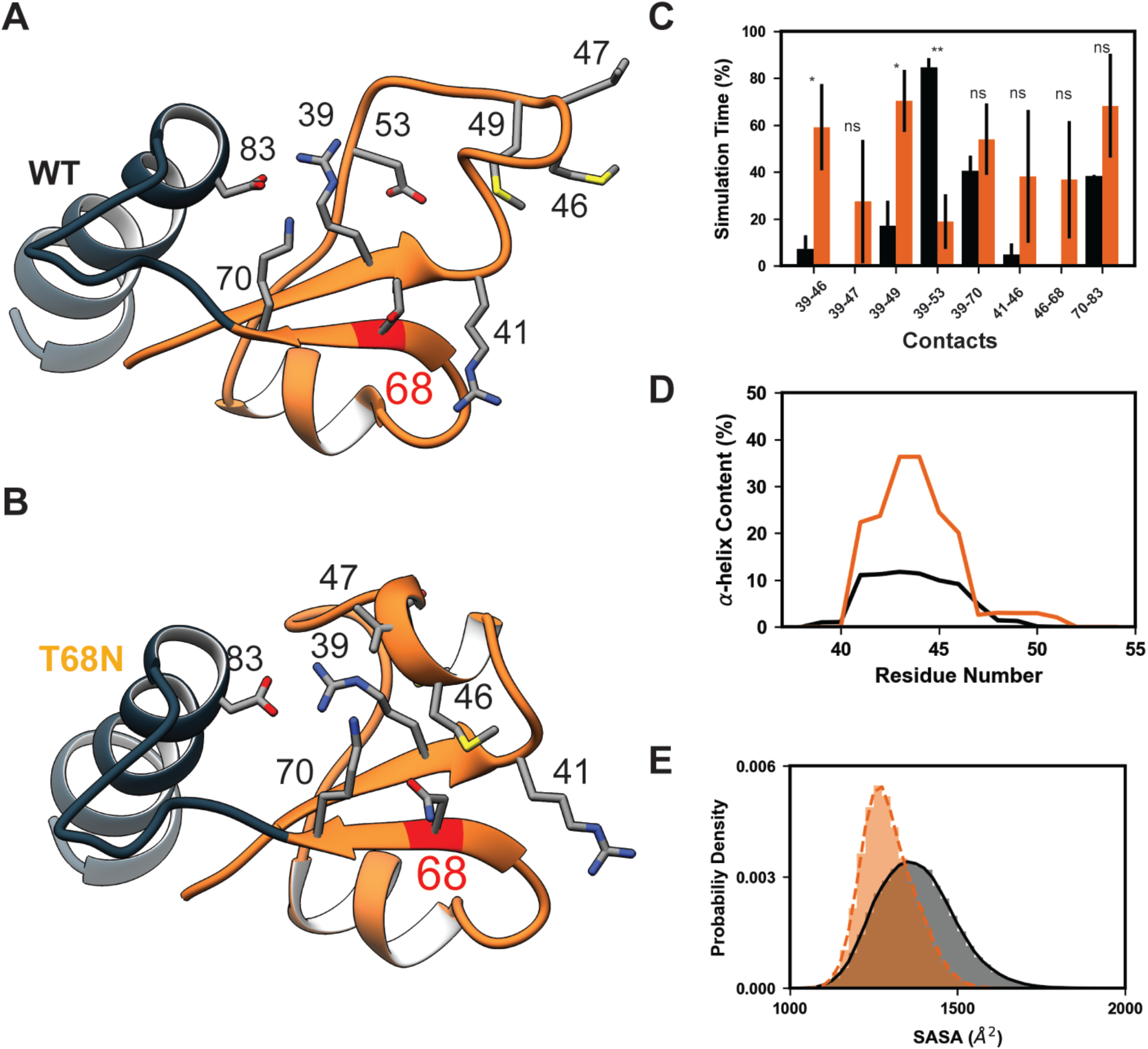
Molecular structures of the globular and filamentous forms of human cardiac actin. (A) Globular (g-) actin monomers are comprised of four subdomains (subdomain 1-4, SD1-4) arranged around the nucleotide binding pocket. The g-actin monomer simulated in this study contains ATP in the binding pocket. The atoms of four residues corresponding to mutation sites examined in this study are shown as spheres: T68 (orange), R185 (magenta), G199 (purple), and R374 (blue). (B) Actin monomers polymerize into protofibrils, which then associate with one another to form filamentous (f-) actin. In this f-actin pentamer, chains A, C and E form one protofibril and chains B and D form the other. The pentamer simulated here has ADP molecules in the nucleotide binding pockets. The location of residue T68 is denoted on chain C.

**Table 2.** C_α_ RMSD values for g-actin monomer simulations.

| **System** | **Run 1** | **Run 2** | **Run 3** | **Average** |
| --- | --- | --- | --- | --- |
| **WT** | 2.7 | 2.5 | 2.5 | **2.6** |
| **p.Thr68Asn** | 2.1 | 2.2 | 1.8 | **2.0** |
| **p.Arg185Trp** | 2.4 | 2.5 | 2.3 | **2.4** |
| **p.Gly199Ser** | 2.7 | 2.5 | 2.1 | **2.4** |
| **p.Arg374Ser** | 2.4 | 2.2 | 2.1 | **2.2** |

### p.Thr68Asn modified the structural organization of subdomain 2 and the D-loop

The RMSD and RMSF data indicated substantial changes in the dynamics of the D-loop in the presence of all four simulated mutations. Therefore, we examined the dynamics in this region in greater detail with an emphasis on the p.Thr68Asn mutation, as T68 is located within SD2 and closest structurally to the D-loop. Mutation of Thr to Asn (p.Thr68Asn) is somewhat conservative: both residues have polar side chains; however, the Asn side chain is long whereas Thr branches at C_b_. The alternate conformations accessible to Asn led to a cascade of changes in amino acid interactions among neighboring residues in SD2 (Figure 4A-C). The changes in contacts affected interactions made by D-loop residues as well as a complex salt bridge formed between residues 39, 70, and 83. p.Thr68Asn increased the extent to which residues in the D-loop formed a-helix secondary structure (Figure 4B, D). p.Thr68Asn decreased the overall solvent accessible surface area (SASA) of residues in the D-loop relative to the WT simulations (Figure 4E). The net effect of the structural changes induced by p.Thr68Asn shifted the structure and dynamic behavior of SD2 such that the mutant SD2 adopted a more compact and less flexible conformation relative to WT.

**Figure 4.**
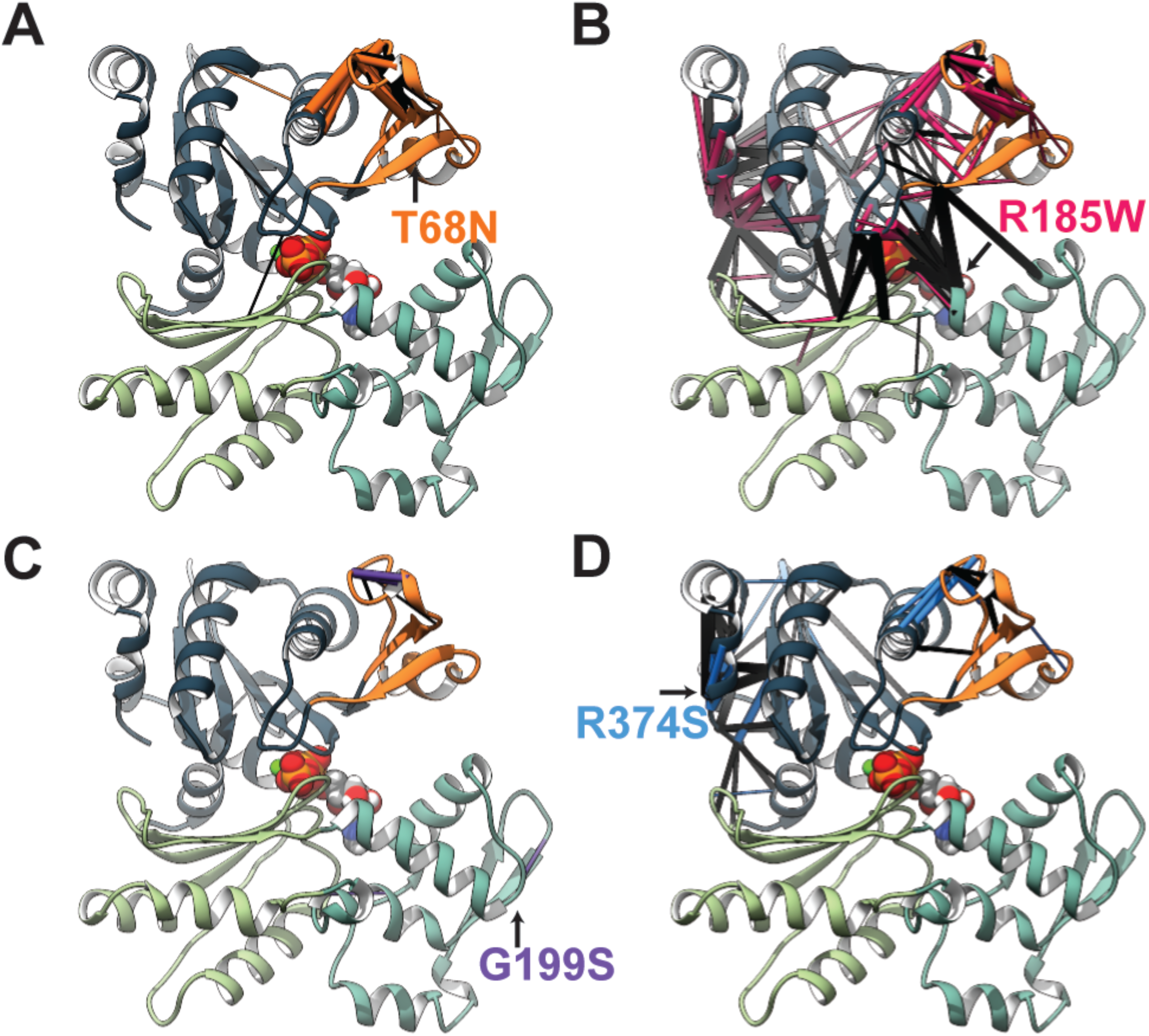
p.Thr68Asn alters the structure and dynamics of g-actin subdomain 2. Comparison of representative MD-derived snapshots of WT (A) and p.Thr68Asn (B) g-actin highlights the structural and dynamical changes induced by p.Thr68Asn (red ribbon). Sidechain atoms of relevant residues are shown and annotated. (C) Bar heights correspond to the fraction of time that select residue pairs spent in contact with one another, averaged over triplicate simulations (error bars correspond to st. dev.). p.Thr68Asn (orange bars) led to shifts in several amino acid interactions relative to WT (black bars). Statistically significant differences between the WT and p.Thr68Asn contact frequencies are denoted (ns: not significant, *: p ≤ 0.05, **: p ≤ 0.01) (D) p.Thr68Asn (orange) increased the α-helix secondary structure content of residues 40-50 within the D-loop relative to WT (black). (E) The p.Thr68Asn mutation (orange) led to a decrease in the solvent accessible surface area of D-loop residues (41-56) relative to WT (black). The histogram shows the SASA probability density of all replicate simulations combined.

These effects were most pronounced for p.Thr68Asn but was also observed for the other mutants. The greater effect of p.Thr68Asn was likely due to its central position in SD2. Altered structure and dynamics among D-loop residues were also observed for the p.Gly199Ser, p.Arg185Trp, and p.Arg374Ser simulations (Figure S4-6). All mutations altered inter-residue interactions formed by D-loop residues and other SD2 residues (Figure S4). All mutations increased the extent to which residues in the D-loop formed α-helix secondary structure in the ensemble average (p.Arg374Ser > p.Arg185Trp > p.Gly199Ser > p.Thr68Asn > WT, Figure S5). However, there was not a statistically meaningful change in the net amount of α-helix formed and this region did form an enduring α-helix in one of the WT simulations. All mutations decreased the average D-loop SASA (WT (1375 Å^2^) > p.Arg374Ser (1349 Å^2^, t-statistic = 0.70, p-value = 0.52) > p.Gly199Ser (1335 Å^2^, t-statistic = 1.47, p-value = 0.21) > p.Arg185Trp (1325 Å^2^, t-statistic = 3.30, p-value = 0.03) > p.Thr68Asn (1294 Å^2^, t-statistic = 3.53, p-value = 0.02), Figure S6). We analyzed statistically significant (Figures S4 and 4; Table S2) changes in residue-residue contact networks to identify structural pathways by which the mutants altered SD2 dynamics (Figure 5, Table S2). Altered residue-residue interactions were only considered in this analysis if there was at least a 10% difference in the average contact time frequency between the WT and mutant simulations. The extent to which the mutations altered residue-residue interaction networks was variable. Disruption was greatest for the p.Arg185Trp mutation and the p.Gly199Ser mutation was the least impactful. p.Thr68Asn modified local residue-residue interactions to affect change in SD2 and the D-loop. p.Arg185Trp, p.Gly199Ser, and p.Arg374Ser instead introduced structural changes that propagated through SD4 and/or SD2 before ultimately altering SD2 structure (Figure 5, SI, Table S2). Though operating through distinct mechanisms, all *ACTC1* mutations simulated in this study led to a common change in the structure and dynamics of SD2 and the D-loop.

**Figure 5.**
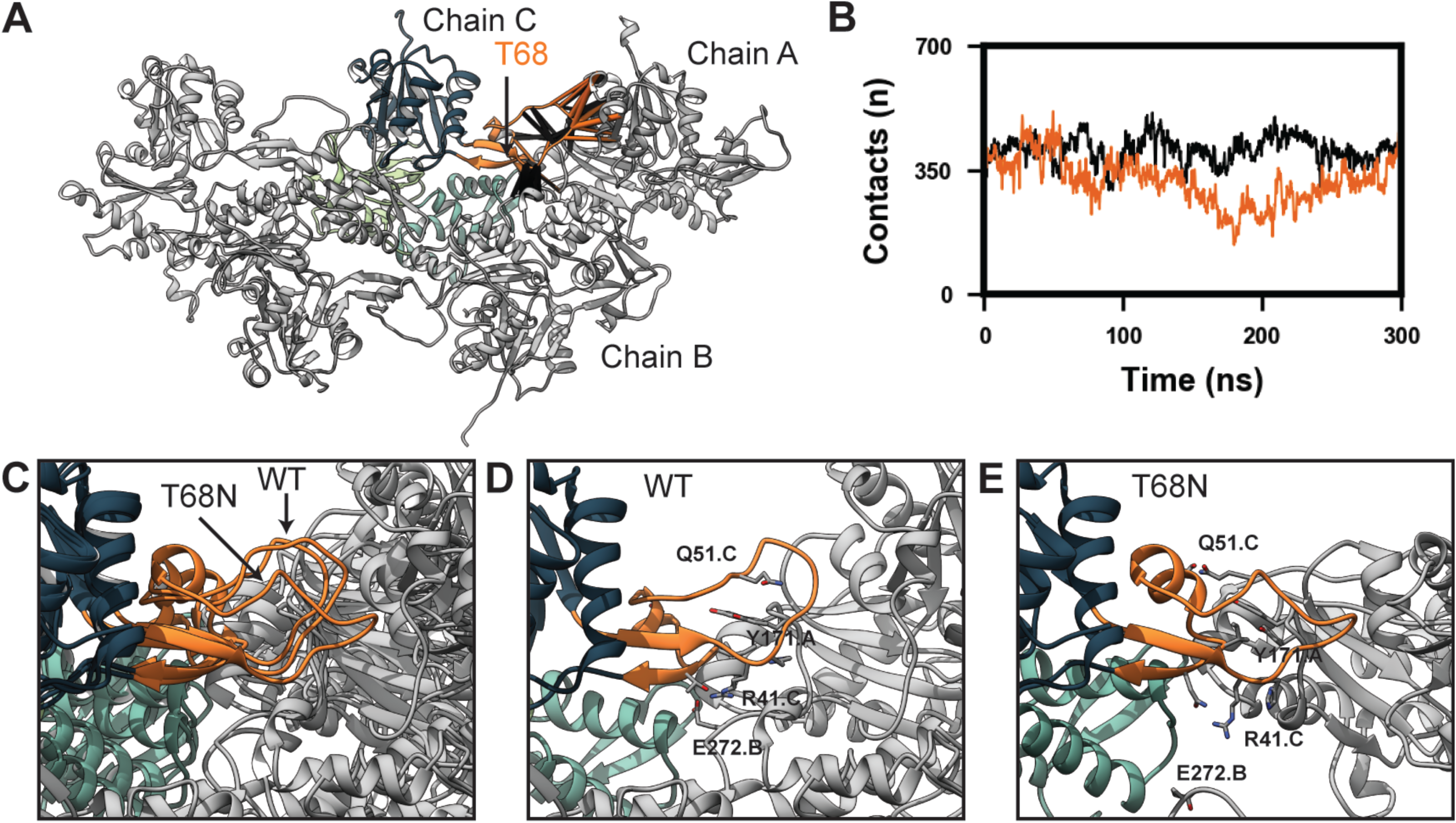
DA-associated mutations lead to structural changes within subdomain 2. The percent simulation time for which residue-residue contacts endured were compared between the four mutant simulations: p.Thr68Asn (A, orange), p.Arg185Trp (B, magenta), p.Gly199Ser (C, purple), p.Arg374Ser (D, blue) and the WT simulations (black). For each mutant-WT comparison, those residue-residue contacts that were present for statistically different percentages of the simulations have been mapped onto the reference crystal structure of g-actin. Contacts that were present more frequently in the WT simulations are denoted by black pipes and contacts present more frequently in the mutant simulations are colored orange, magenta, purple, or blue. The thickness of the pipes corresponds to the difference in percent simulation time that the contact was present between the WT and mutant simulations (larger pipes indicate a contact was observed more frequently). Although the mutations were distributed throughout the structure, they all led to statistically significant (see Table S2 for test statistics) changes in the structure of subdomain 2 (orange ribbons).

### p.Thr68Asn modified interactions between actin subunits in f-actin

In our g-actin simulations, all mutations led to similar structural changes in SD2 and the D-loop. To make predictions about the effects of altered SD2 structure/dynamics in actin filaments, we performed MD simulations of WT and p.Thr68Asn cardiac f-actin pentamers (Figure 3B). Simulation performance rapidly decreases with the number of atoms; therefore, we simulated pentamers as a simplified proxy for actin filaments and only analyzed the dynamics of the central chain (chain C) to avoid end effects. f-actin filaments are composed of two proto-fibrils, each of which contains monomers arranged such that SD3 of one monomer is inserted into the cleft between SD2 and SD4 of the succeeding monomer (moving from the – to + end). Two protofilaments twist around one another and the face of the monomer containing the mutations (the front facing side in Figure 3B) is buried. SD2 is a critical structural component of actin filaments: it forms interactions between actins within a single protofilament (intra-filament) and between monomers of different protofilaments (inter-filament). For example, in the cryoEM structure, the D-loop of one monomer encircles Y171 of the succeeding monomer in the same protofilament (in our model, the D-loop of chain C loops around Y171 of chain A). Additionally, R41 forms a salt bridge with E272 of a monomer in the opposite protofilament. In GaMD simulations of f-actin, the p.Thr68Asn resulted in a shift in the structure and dynamics of SD2 and the D-loop (Figure 6). The mutation resulted in a change in residue-residue interactions made by the D-loop (Figure 6A) and reduced the number of contacts made between SD2 of chain C and atoms in other actin subunits (Figure 6B). As was observed in the g-actin simulations, p.Thr68Asn promoted a more compact conformation of the D-loop in f-actin (Figure 6C). The more compact loop conformation reduced interactions between the D-loop of chain C and Y171 of chain A and also eliminated the salt bridge formed between R41 of chain C and E272 of chain B (Figure 6D,E).

**Figure 6.** p.Thr68Asn alters inter-chain interactions made by subdomain 2 in f-actin. Residue-residue interactions formed between SD2 of chain C and chains A and B were analyzed in the GaMD simulations of WT and p.Thr68Asn f-actin. (A) p.Thr68Asn led to statistically significant differences in residue-residue contacts formed by SD2 of chain C (denoted by pipes as in Figure 5). Differences were found in contacts formed between SD2 of chain C and SD1 of chain A, as well as in contacts formed between SD2 of chain C and the SD3-SD4 linker of chain B. (B) The total number of atom-atom interactions formed between SD2 of chain C and all atoms in chains A and B were monitored in the WT and p.Thr68Asn GaMD simulations. Relative to the WT simulation (black) the p.Thr68Asn simulation (orange) had fewer inter-chain contacts involving chain CSD2. (C) In the reference cryoEM structure and WT simulation, the D-loop of SD2 in chain C fits into a pocket formed by SD1 and SD3 of chain A. The p.Thr68Asn simulations instead sampled non-native conformations in which the D-loop exited this binding pocket. In the reference cryoEM structure and the WT simulation (D) the D-loop of chain C is stabilized via a network of hydrophobic interactions formed with Y171 of chain A as well as a hydrogen bond network involving Arg 41 (chain C), Thr 68 (chain C), and Glu 272 (chain B). These interactions were disrupted in the p.Thr68Asn simulation (E).

## Discussion

We identified five unrelated families in which a total of eight individuals have heterozygous, rare, pathogenic variants in *ACTC1* and share similar phenotypic effects including multiple congenital contractures, neck pterygia, scoliosis, and congenital heart defects/cardiomyopathy. This pattern of clinical findings appears to represent an autosomal dominant disorder distinct from previously reported Mendelian conditions due to *ACTC1* variants that are characterized by cardiac abnormalities including autosomal dominant atrial septal defects [MIM 612794],^44^ dilated cardiomyopathy [MIM 613424],^45^ hypertrophic cardiomyopathy [MIM 612098],^46^ and left ventricular noncompaction [MIM 613424]^47^). MD simulations demonstrate that all four variants (p.Thr68Asn, p.Arg185Trp, G19S, and p.Arg374Ser) disrupt the native structure of the regions of actin most associated with protein-protein interactions (subdomain 2, or SD2, and the D-loop), impeding interactions between actin and its binding partners, including other actins during thin filament assembly. Additionally, the altered D-loop structure is predicted to increase structural disorder within thin filaments, resulting in ‘stretchier’ thin filaments that may contract more slowly, require greater loads to extend, have weakened force production, and/or have slower rates of force production. Structural perturbations to the D-loop are known to affect thin filament stiffness.^48, 49^ Thus, while the genetic basis of this DA condition is unique compared to other DAs, the underlying mechanisms may be similar if not identical.^9, 15, 50–52^

All of the pathogenic *ACTC1* variants (n = 87) reported to date (Table S1), with the exception of p.Arg185Trp that we also identified in Family D, were found in persons noted only to have abnormalities of the heart. While it is possible that congenital contractures have been overlooked in these families, this seems like an unlikely explanation for all or even most families. Alternatively, there may be a biological explanation(s) for this observation, none of which are mutually exclusive. First, none of the residues perturbed in the families we identified, except for Arg185 which was previously found in a person with congenital contractures, have been reported previously. So, the distribution of phenotypic effects associated with these genotypes has been, to date, unknown. Second, substitutions of each of these residues in ACTC1 increases disorder of actin SD2 and D-loop interactions, and these perturbations could be a unique consequence of contracture-associated variants. Third, the presence of a pathogenic *ACTC1* variant may be necessary but not sufficient for the development of congenital contractures. In other words, skeletal muscle might be affected only in the presence of genetic modifier(s). We verified the absence of additional rare coding *ACTC1* or *ACTA1* variants but could not exclude the presence of structural variants and/or variants in non-coding regulatory elements that might alter expression of *ACTC1* or *ACTA1*.

The observation that rare genotypes in ACTC1 underlie both cardiac and skeletal abnormalities is not without precedent. ACTC1 and ACTA1 are highly homologous, differing by only four amino acids (Figure 2), and both actins are expressed in skeletal and cardiac muscle.^53–56^ During fetal development, ACTC1 is the predominant actin, as measured by protein expression, in both skeletal and cardiac muscle,.^55^ It is downregulated starting around 27-28 weeks of fetal development and continues to decline until ∼6 months of age when it accounts for ∼5% of total actin.^55^ In adult skeletal muscle ACTC1 and ACTA1 account for ∼5% and 95% of actin, respectively,^55^ and ACTC1 accounts for ∼80% of actin in adult cardiac muscle.^53^ These differences in spatial and temporal expression are considered explanations of the exclusive association of skeletal muscle abnormalities (i.e., nemaline myopathy [MIM 161800], actin accumulation myopathy [MIM 161800], congenital fiber-type disproportion [MIM 255310], intranuclear rod myopathy [MIM 161800], etc.) with *ACTA1* mutations and cardiac abnormalities (autosomal dominant atrial septal defects [MIM 612794],^44^ dilated [MIM 613424]^45^ and hypertrophic [MIM 612098]^46^ cardiomyopathy, and left ventricular noncompaction [MIM 613424]^47^) with mutations in *ACTC1*. Of the hundreds of individuals described with *ACTA1*-associated myopathy, only twelve (nine unique variants)^57–65^ have been reported to also have a cardiac abnormality (Table S1), either in conjunction with a skeletal myopathy (N=10) or alone (N=2). Rare variants in *ACTC1* resulting in congenital contractures in a small fraction of persons with *ACTC1* variants appears to be the corollary.

Mutations in *ACTC1* result in skeletal muscle contractures even though ACTC1 accounts for only ∼5% of total actin in adult skeletal muscle.^53^ The most likely explanation is that the skeletal muscle contractures originate during fetal development when ACTC1 is the predominant source of sarcomeric actin, and replacement of most skeletal muscle actin with wildtype ACTA1 during infancy is insufficient to correct the abnormality. However, it is possible that *ACTC1* has a previously unknown function in skeletal muscle biology, or that mutant ACTC1 protein interferes with the function of actin encoded by *ACTA1*. Testing this hypothesis will require further functional characterization of these variants.

The MD simulations are limited by several factors. First and foremost, the method used to introduce mutations assumes that the mutant constructs can access WT-like conformations and the timescale along which they transition from a WT-like ensemble to a mutant ensemble are not known. Second, our simulations have probed isolated states of g-actin and f-actin and cannot directly describe effects that the mutations have on interactions between g-actin and its binding partners nor between f-actin and the rest of the contractile machinery present in sarcomeres. Nevertheless, these simulations provide predictions about the functional consequences of DA-associated mutations in *ACTC1* generate hypotheses on disease mechanisms and provide guidance for future studies such as investigation of whether the mutations result in impaired filament assembly or impaired filament mechanics.

In summary, we identified five unrelated families with heterozygous pathogenic variants in *ACTC1* resulting in multiple congenital contractures, webbed neck, scoliosis, short stature, and distinctive facial features as well as cardiac abnormalities including atrial and ventricular septal defects, left ventricular noncompaction, cardiomyopathy. This appears to be a novel Mendelian condition due to pathogenic variants in a gene known to underlie conditions characterized only by cardiac defects. Our findings suggest both that persons with multiple congenital contractures should be tested for pathogenic variants in *ACTC1* and persons with contractures and pathogenic variants in *ACTC1* should undergo cardiac evaluation for both structural and functional abnormalities.

## Data Availability

Sequence data for Families A is in dbGaP under accession number phs000693 and B and C will be available in the AnVIL under accession number phs003047 pending the first public release of the GREGoR dataset.

## Supplemental Information

Supplemental Information include 6 figures, 1 table, and detailed description of MD simulation results.

## Data and Code Availability

Sequence data for Families A is in dbGaP under accession number phs000693 and B and C will be available in the AnVIL under accession number phs003047 pending the first public release of the GREGoR dataset. Please contact the corresponding author M.J.B for further information.

## Acknowledgements

We thank the families for their participation and support. Sequencing and data analysis were provided by the University of Washington Center for Rare Disease Research (UW-CRDR) with support from NHGRI grants U01 HG011744, UM1 HG006493 and U24 HG011746. Protein simulations were performed with resources provided by the UW Center for Translational Muscle Research (PI, Regnier), which is supported by the National Institute of Arthritis and Musculoskeletal and Skin Diseases of the National Institutes of Health under Award Number P30AR074990. Additional funding support for MR and MCC was provided by NIH RM1 GM131981. This work was also funded by the National Institute of Child Health and Human Development (4R01HD048895 to M.J.B.), and a Cardiovascular Research Training Grant from the National Heart, Lung, and Blood Institute (T32HL007828 to M.C.C.). The content is solely the responsibility of the authors and does not necessarily represent the official views of the National Institutes of Health.

## Web resources

The URLs for data presented herein are as follows: MyGene2: https://mygene2.org

Geno2MP: https://geno2mp.gs.washington.edu/Geno2MP/ gnomAD: http://gnomad.broadinstitute.org

Human Genome Variation: http://www.hgvs.org/mutnomen/

Online Mendelian Inheritance in Man (OMIM): http://www.omim.org/

