## Supplemental methods, figures, and table legends for "Variants in *ACTC1* underlie distal arthrogryposis accompanied by congenital heart defects"

**Supplemental Materials and Methods**

**Table of Contents**

1. Summary of structural effects of R185W, G199S, and R374S
2. Figure S1. Pedigrees of families with pathogenic variants in *ACTC1* resulting in distal arthrogryposis
3. Figure S2. Histochemistry and immunohistochemistry from muscle biopsy of paravertebral muscles in proband of Family D.
4. Figure S3. Mutation-associated changes in C_α_ RMSF of g-actin.
5. Figure S4. Mutation-associated changes in D-loop – subdomain 2 contacts.
6. Figure S5. Mutation-associated changes in D-loop secondary structure.
7. Figure S6. Mutation-associated changes in D-loop solvent accessible surface area.
8. Table S1. *ACTC1* and *ACTA1* missense variants reported as pathogenic in individuals with phenotypes involving cardiac muscle, skeletal muscle, or both. (provided as a separate Excel spreadsheet)
9. Table S2. Reported residue-residue pair interaction frequencies that differed among the WT and mutant simulations. (provided as a separate Excel spreadsheet and in this document)

**Summary of structural effects of R185W, G199S, and R374S**

R185Y: In the WT simulations, R185 formed transient electrostatic interactions with S16, G17, E74, and D159. Additionally the Y71 side chain formed a large number of atom-atom contacts with R185 by stacking against its guanidino group. Due to these interactions, R185 stabilizes g-actin structure in the vicinity of the ATP pocket and forms a structural linker between all four subdomains. Mutation of R185 to Trp eliminated these stabilizing interactions; instead, W185 was primarily positioned in the cleft between SD2 and SD4 and interacted primarily with other SD4 residues. This resulted in a large-scale reorganization of inter-residue contacts spanning all 4 subdomains and an increase in the SD2-SD4 cleft distance.

G199S: In the WT simulations, G199 adopts backbone dihedral angles that are unfavorable for most amino acids and accommodates a transition from α-helix structure in residues 184-198 to coil structure in residues 199 – 204. Mutation to S199 resulted in a shift in the backbone conformation of residue 199 to angles within the lower left quadrant of Ramachandran space, a decrease in a-helix structure in residues 197-198, and a shift in the structure of the loop spanning residues 199 to 204. This change resulted in few statistically significant changes in inter-residue interactions but did modify the breathing motion between subdomains 2 and 4.

R374S: In the WT simulations, R374 formed transient salt bridges with E363 and E366. Each of these residues are located on the surface of SD1. These salt bridges were lost in the R374S simulations. Simultaneously there was a reorganization of inter-residue interactions in SD1 as well as an increase in residue-residue interactions between an α-helix of SD1 (residues 81-96) and residues in SD2.

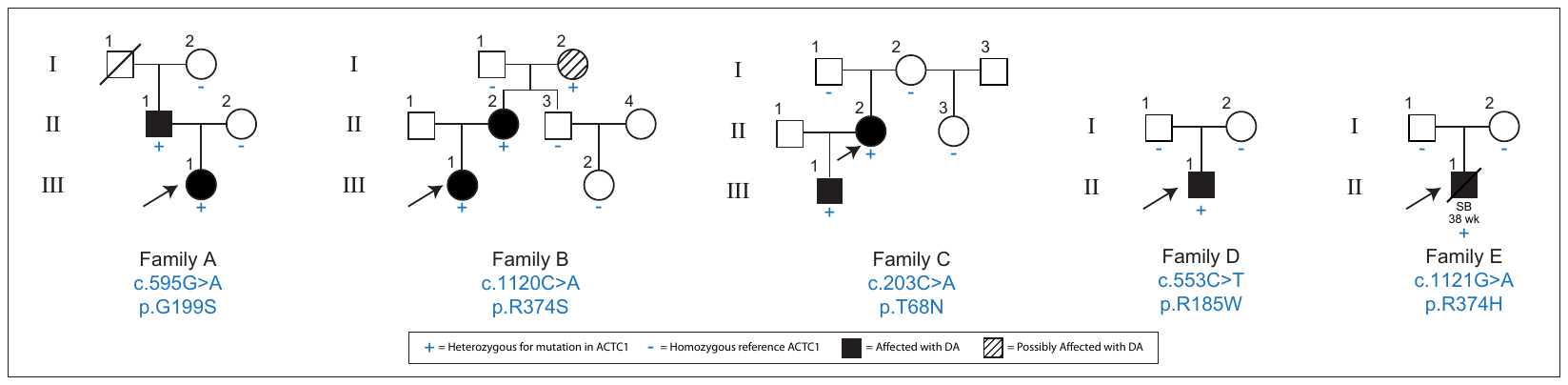
**Figure S1.** Pedigrees of families with pathogenic variants in *ACTC1* resulting in distal arthrogryposis.

**Figure S2. Histochemistry and immunohistochemistry from muscle biopsy of paravertebral muscles in proband of Family D.** (A) Scattered areas of round muscle fibers found in H&E [hematoxylin and eosin] staining. (B) ATPase (pH4.2) shows type I fiber predominance that could be seen in paravertebral muscles. (C) No ragged red fibers or rod-like inclusions are seen with modified Gomori trichrome staining however scattered nemaline bodies were found in (D) electronic microscopy.

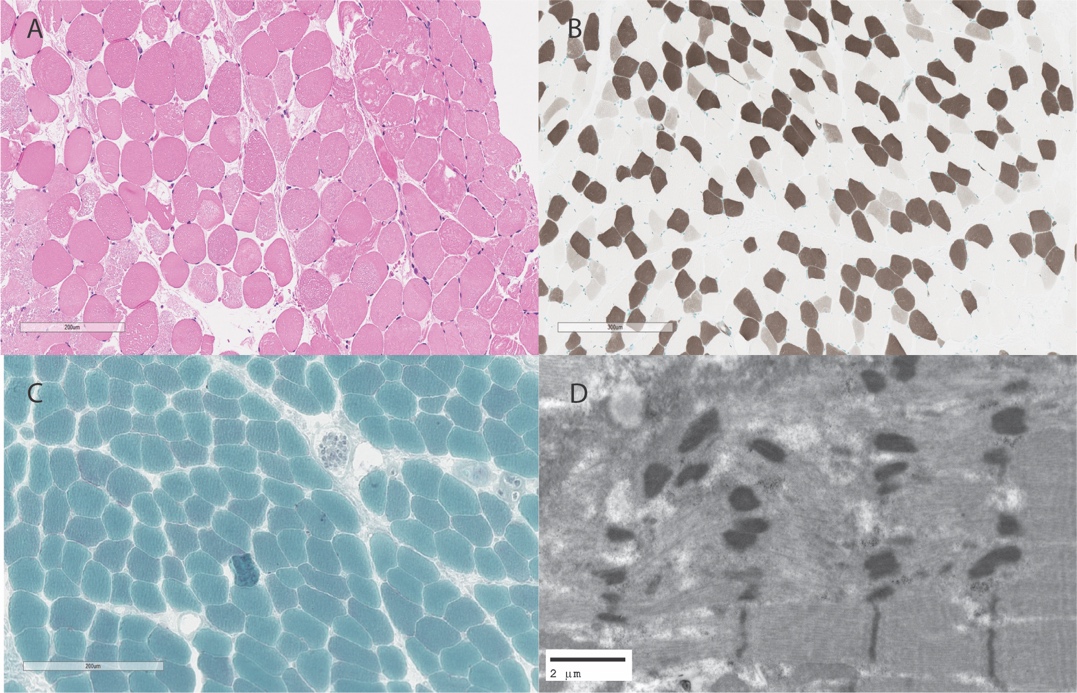

**Figure S3. Mutation-associated changes in C_α_ RMSF of g-actin.** In MD simulations, C_α_ RMSF measures the root-mean-squared fluctuation of the C_α_ atoms about their average positions. RMSF values were calculated after alignment of the g-actin trajectories to their average coordinates. Average RMSF values are reported per residue (left column) for the WT (a), T68N (b), R185W (c), G199S (d), and R374S (e)., simulations. Points correspond to the average value and shaded regions denote the standard deviation. Vertical red lines denote mutation sites. The difference in the average RMSF relative to the WT simulation of the T68N (b), R185W (c), G199S (d), and R374S (e) mutations are plotted in the right column. Positive values indicate greater RMSF in WT and negative values indicate greater RMSF in the mutants. Vertical red lines denote mutation sites. Red points denote residues with statistically significant (p < 0.05) differences in the RMSF between the WT and mutant simulations.

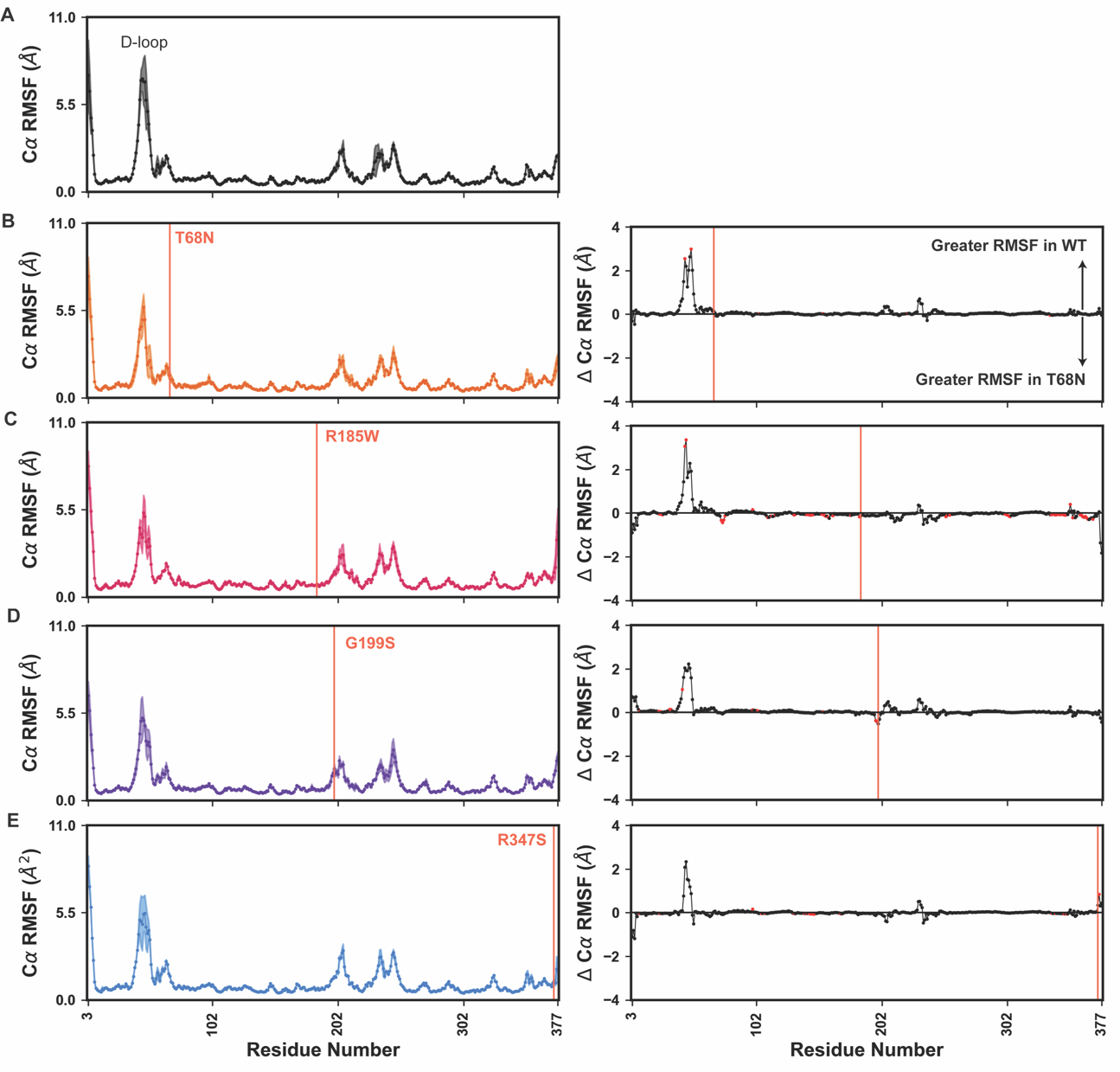

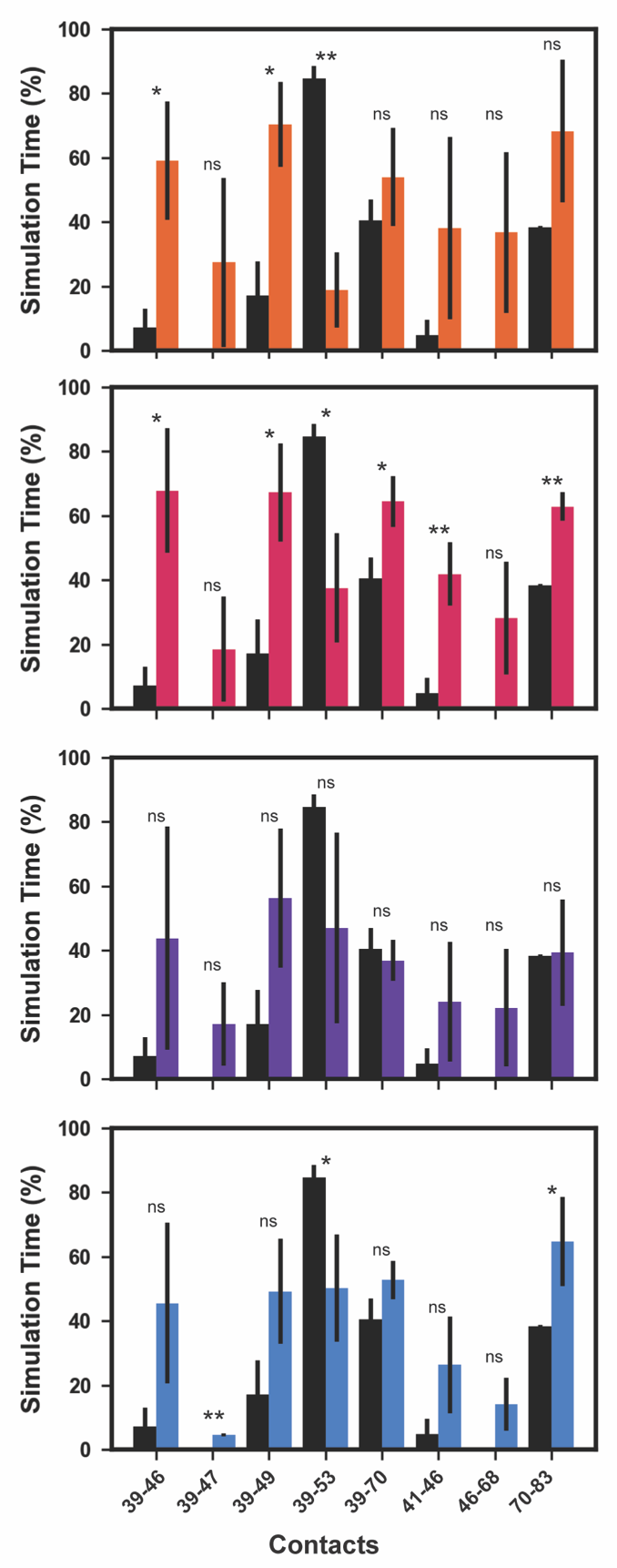
**Figure S4. Mutation-associated changes in D-loop – subdomain 2 contacts.** We calculated the average percent simulation time for which select residue-residue interactions were present in the WT (black), T68N (orange), R185W (magenta), G199S (purple), and R374S (blue) simulations. Error bars denote standard deviation. Statistically significant differences between the WT and T68N contact frequencies are denoted (ns: not significant, *: p ≤ 0.05, **: p ≤ 0.01)

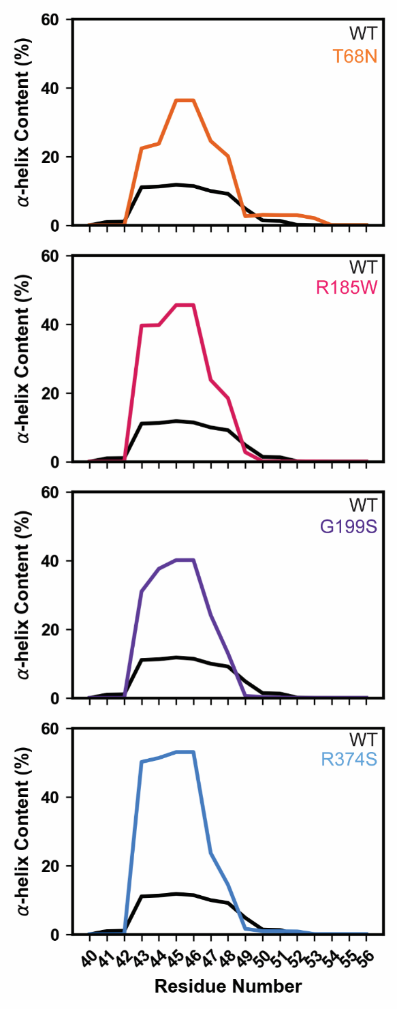
**Figure S5. Mutation-associated changes in D-loop secondary structure.** For each residue in the D-loop (# 40 – 56), we calculated the average percent simulation time for which the residue adopted α-helix secondary structure. The α-helix content per residue is shown for T68N (orange), R185W (magenta), G199S (purple), and R374S (blue) versus the WT (grey) simulation. All mutations in this study resulted in an increase in α-helix content in the D-loop.

**Figure S6. Mutation-associated changes in D-loop solvent accessible surface area.** Histograms of the solvent accessible surface area aggregated over all replicate simulations are plotted for T68N (orange), R185W (magenta), G199S (purple), and R374S (blue) versus the WT (grey) distribution. All mutations in this study resulted in a decrease in the SASA of D-loop residues.

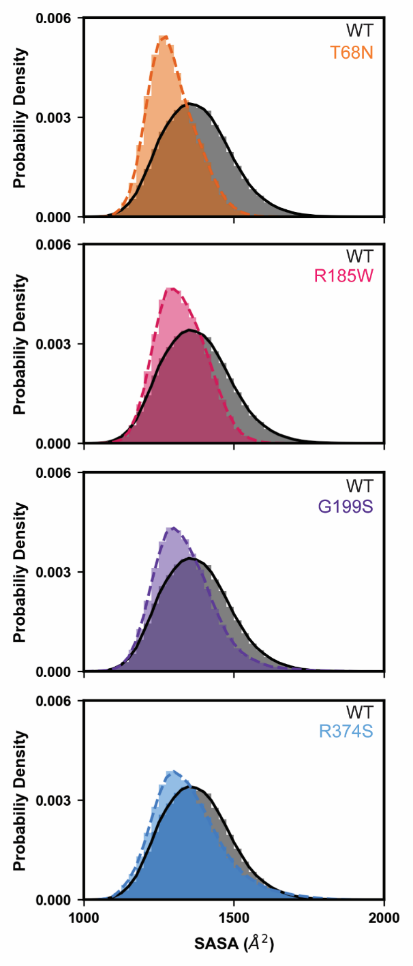

**Table S1. *ACTC1* and *ACTA1* missense variants reported as pathogenic in individuals with phenotypes involving cardiac muscle, skeletal muscle, or both.** This table contains all missense variants in *ACTA1* or *ACTC1* that were reported as Likely Pathogenic or Pathogenic in ClinVar; Damaging in HGMD, and/or non-synonymous in the *ACTC1* or *ACTA1* LOVD Locus Specific Databases. ACTA1 and ACTC1 are both 377 residues long and differ at only 4 sites (4, 5, 301, 360). Each variant is listed alongside the reported phenotype (one row per affected individual/database entry), whether the affected individual had skeletal muscle or cardiac muscle findings, the Pubmed PMID of any associated publication and/or submitter. The residue numbering listed for ACTA1 variants was updated to current (2022) numbering if necessary.

Table S1 is provided as an Excel spreadsheet.

**Table S2. Reported residue-residue pair interaction frequencies that differed among the WT and mutant simulations.** This table enumerates the average percent simulation time for which select residue-residue pairs formed inter-residue interactions in the WT and mutant MD simulations. Altered residue-residue interactions are only reported if there was at least a 10% difference in the average contact time frequency between the WT and mutant simulations and a statistically significant difference in the average WT and mutant interaction frequencies.

Table S2 is also provided as an Excel spreadsheet.

| **Mutation** | **Residue I** | **Residue J** | **Contact Frequency**  **(WT, %)** | **Contact Frequency**  **(Mutant, %)** | **T-test**  **Statistic** | **P-Value** |
| --- | --- | --- | --- | --- | --- | --- |
| **T68N** | 3 | 103 | 26 | 4 | 2.834 | 0.047 |
| **T68N** | 14 | 121 | 92 | 80 | 5.711 | 0.005 |
| **T68N** | 39 | 46 | 7 | 59 | -3.79 | 0.019 |
| **T68N** | 39 | 49 | 17 | 70 | -4.47 | 0.011 |
| **T68N** | 39 | 52 | 42 | 8 | 3.805 | 0.019 |
| **T68N** | 39 | 53 | 85 | 19 | 7.6 | 0.002 |
| **T68N** | 40 | 51 | 33 | 0 | 3.085 | 0.037 |
| **T68N** | 41 | 51 | 13 | 0 | 5.527 | 0.005 |
| **T68N** | 41 | 65 | 74 | 87 | -3.475 | 0.025 |
| **T68N** | 41 | 67 | 82 | 94 | -3.185 | 0.033 |
| **T68N** | 42 | 51 | 23 | 0 | 3.551 | 0.024 |
| **T68N** | 43 | 51 | 47 | 3 | 4.354 | 0.012 |
| **T68N** | 44 | 51 | 27 | 0 | 4.167 | 0.014 |
| **T68N** | 45 | 49 | 24 | 76 | -2.977 | 0.041 |
| **T68N** | 46 | 51 | 24 | 5 | 5.234 | 0.006 |
| **T68N** | 47 | 51 | 33 | 5 | 4.41 | 0.012 |
| **T68N** | 50 | 54 | 16 | 87 | -5.7 | 0.005 |
| **T68N** | 51 | 54 | 13 | 88 | -8.014 | 0.001 |
| **T68N** | 51 | 82 | 8 | 88 | -7.869 | 0.001 |
| **T68N** | 51 | 83 | 6 | 59 | -7.649 | 0.002 |
| **T68N** | 51 | 86 | 6 | 84 | -7.427 | 0.002 |
| **T68N** | 52 | 86 | 22 | 88 | -7.062 | 0.002 |
| **T68N** | 75 | 163 | 85 | 73 | 2.923 | 0.043 |
| **T68N** | 92 | 131 | 8 | 18 | -2.846 | 0.047 |
| **R185W** | 7 | 355 | 64 | 18 | 4.728 | 0.009 |
| **R185W** | 8 | 355 | 38 | 2 | 3.4 | 0.027 |
| **R185W** | 9 | 355 | 57 | 4 | 4.689 | 0.009 |
| **R185W** | 10 | 92 | 84 | 96 | -5.449 | 0.006 |
| **R185W** | 10 | 358 | 84 | 67 | 7.614 | 0.002 |
| **R185W** | 11 | 105 | 40 | 26 | 2.937 | 0.043 |
| **R185W** | 12 | 91 | 54 | 81 | -6.932 | 0.002 |
| **R185W** | 12 | 92 | 76 | 50 | 5.995 | 0.004 |
| **R185W** | 12 | 105 | 56 | 37 | 4.637 | 0.01 |
| **R185W** | 14 | 19 | 86 | 72 | 7.18 | 0.002 |
| **R185W** | 14 | 76 | 8 | 42 | -3.143 | 0.035 |
| **R185W** | 16 | 74 | 6 | 79 | -5.433 | 0.006 |
| **R185W** | 16 | 75 | 12 | 85 | -5.969 | 0.004 |
| **R185W** | 16 | 77 | 1 | 19 | -3.258 | 0.031 |
| **R185W** | 16 | 185 | 100 | 0 | 726.152 | 0 |
| **R185W** | 17 | 36 | 25 | 13 | 3.079 | 0.037 |
| **R185W** | 17 | 185 | 99 | 0 | 295.7 | 0 |
| **R185W** | 19 | 34 | 72 | 85 | -3.348 | 0.029 |
| **R185W** | 19 | 84 | 73 | 47 | 3.512 | 0.025 |
| **R185W** | 19 | 88 | 43 | 12 | 10.62 | 0 |
| **R185W** | 21 | 92 | 22 | 8 | 7.167 | 0.002 |
| **R185W** | 23 | 96 | 74 | 88 | -5.732 | 0.005 |
| **R185W** | 23 | 103 | 73 | 50 | 3.082 | 0.037 |
| **R185W** | 25 | 347 | 51 | 39 | 4.542 | 0.01 |
| **R185W** | 31 | 91 | 40 | 21 | 5.952 | 0.004 |
| **R185W** | 32 | 58 | 6 | 27 | -6.528 | 0.003 |
| **R185W** | 32 | 95 | 60 | 36 | 6.647 | 0.003 |
| **R185W** | 34 | 71 | 1 | 15 | -7.819 | 0.001 |
| **R185W** | 34 | 87 | 88 | 68 | 3.949 | 0.017 |
| **R185W** | 35 | 73 | 63 | 89 | -3.77 | 0.02 |
| **R185W** | 35 | 84 | 24 | 9 | 3.931 | 0.017 |
| **R185W** | 37 | 86 | 49 | 87 | -6.088 | 0.004 |
| **R185W** | 37 | 87 | 87 | 97 | -3.547 | 0.024 |
| **R185W** | 38 | 56 | 67 | 80 | -3.033 | 0.039 |
| **R185W** | 39 | 46 | 7 | 68 | -4.233 | 0.013 |
| **R185W** | 39 | 49 | 17 | 67 | -3.824 | 0.019 |
| **R185W** | 39 | 52 | 42 | 4 | 5.082 | 0.007 |
| **R185W** | 39 | 53 | 85 | 38 | 3.823 | 0.019 |
| **R185W** | 39 | 70 | 40 | 64 | -3.301 | 0.03 |
| **R185W** | 40 | 45 | 21 | 90 | -4.736 | 0.009 |
| **R185W** | 40 | 46 | 23 | 91 | -5.749 | 0.005 |
| **R185W** | 41 | 46 | 5 | 42 | -4.787 | 0.009 |
| **R185W** | 41 | 64 | 50 | 66 | -7.799 | 0.001 |
| **R185W** | 41 | 65 | 74 | 88 | -3.734 | 0.02 |
| **R185W** | 41 | 67 | 82 | 95 | -3.446 | 0.026 |
| **R185W** | 42 | 45 | 58 | 100 | -5.57 | 0.005 |
| **R185W** | 42 | 48 | 14 | 0 | 3.239 | 0.032 |
| **R185W** | 43 | 51 | 47 | 9 | 2.922 | 0.043 |
| **R185W** | 44 | 51 | 27 | 2 | 3.709 | 0.021 |
| **R185W** | 45 | 49 | 24 | 74 | -2.879 | 0.045 |
| **R185W** | 46 | 51 | 24 | 9 | 3.073 | 0.037 |
| **R185W** | 47 | 51 | 33 | 2 | 6.377 | 0.003 |
| **R185W** | 48 | 51 | 39 | 0 | 11.718 | 0 |
| **R185W** | 50 | 54 | 16 | 68 | -3.051 | 0.038 |
| **R185W** | 51 | 54 | 13 | 67 | -3.625 | 0.022 |
| **R185W** | 51 | 82 | 8 | 48 | -4.509 | 0.011 |
| **R185W** | 51 | 85 | 1 | 30 | -2.847 | 0.047 |
| **R185W** | 51 | 86 | 6 | 71 | -4.372 | 0.012 |
| **R185W** | 52 | 85 | 2 | 18 | -5.681 | 0.005 |
| **R185W** | 56 | 83 | 35 | 7 | 4.063 | 0.015 |
| **R185W** | 56 | 90 | 89 | 99 | -2.79 | 0.049 |
| **R185W** | 57 | 90 | 13 | 37 | -2.934 | 0.043 |
| **R185W** | 61 | 71 | 9 | 29 | -7.417 | 0.002 |
| **R185W** | 70 | 83 | 38 | 63 | -7.659 | 0.002 |
| **R185W** | 71 | 80 | 44 | 25 | 3.283 | 0.03 |
| **R185W** | 71 | 159 | 67 | 24 | 9.272 | 0.001 |
| **R185W** | 71 | 185 | 98 | 18 | 4.678 | 0.009 |
| **R185W** | 71 | 208 | 54 | 1 | 5.683 | 0.005 |
| **R185W** | 73 | 185 | 22 | 0 | 6.856 | 0.002 |
| **R185W** | 74 | 185 | 20 | 0 | 8.777 | 0.001 |
| **R185W** | 75 | 163 | 85 | 38 | 6.651 | 0.003 |
| **R185W** | 75 | 179 | 98 | 75 | 27.142 | 0 |
| **R185W** | 75 | 181 | 78 | 19 | 6.857 | 0.002 |
| **R185W** | 76 | 161 | 81 | 12 | 5.399 | 0.006 |
| **R185W** | 76 | 163 | 45 | 3 | 9.229 | 0.001 |
| **R185W** | 87 | 92 | 22 | 11 | 7.734 | 0.002 |
| **R185W** | 88 | 93 | 74 | 62 | 5.318 | 0.006 |
| **R185W** | 92 | 105 | 81 | 94 | -5.211 | 0.006 |
| **R185W** | 93 | 128 | 50 | 33 | 3.715 | 0.021 |
| **R185W** | 100 | 104 | 39 | 20 | 5.873 | 0.004 |
| **R185W** | 100 | 125 | 19 | 6 | 24.044 | 0 |
| **R185W** | 104 | 355 | 32 | 3 | 3.012 | 0.039 |
| **R185W** | 106 | 136 | 86 | 60 | 7.426 | 0.002 |
| **R185W** | 106 | 137 | 90 | 72 | 5.361 | 0.006 |
| **R185W** | 106 | 348 | 49 | 87 | -5.507 | 0.005 |
| **R185W** | 106 | 354 | 85 | 51 | 3.307 | 0.03 |
| **R185W** | 109 | 377 | 5 | 43 | -2.974 | 0.041 |
| **R185W** | 110 | 163 | 78 | 19 | 4.076 | 0.015 |
| **R185W** | 118 | 376 | 0 | 53 | -5.678 | 0.005 |
| **R185W** | 118 | 377 | 0 | 55 | -5.725 | 0.005 |
| **R185W** | 134 | 357 | 18 | 52 | -4.789 | 0.009 |
| **R185W** | 135 | 348 | 10 | 50 | -7.664 | 0.002 |
| **R185W** | 135 | 372 | 56 | 3 | 2.792 | 0.049 |
| **R185W** | 135 | 374 | 61 | 1 | 3.531 | 0.024 |
| **R185W** | 136 | 376 | 1 | 64 | -10.241 | 0.001 |
| **R185W** | 137 | 348 | 22 | 47 | -5.38 | 0.006 |
| **R185W** | 137 | 376 | 31 | 1 | 3.251 | 0.031 |
| **R185W** | 138 | 377 | 7 | 47 | -2.838 | 0.047 |
| **R185W** | 140 | 156 | 76 | 61 | 4.604 | 0.01 |
| **R185W** | 140 | 341 | 67 | 46 | 2.932 | 0.043 |
| **R185W** | 141 | 376 | 18 | 0 | 3.437 | 0.026 |
| **R185W** | 141 | 377 | 87 | 23 | 13.812 | 0 |
| **R185W** | 145 | 170 | 11 | 32 | -3.201 | 0.033 |
| **R185W** | 145 | 377 | 72 | 16 | 5.403 | 0.006 |
| **R185W** | 150 | 377 | 27 | 4 | 3.163 | 0.034 |
| **R185W** | 158 | 302 | 31 | 6 | 4.475 | 0.011 |
| **R185W** | 159 | 186 | 79 | 30 | 3.04 | 0.038 |
| **R185W** | 160 | 185 | 96 | 22 | 23.03 | 0 |
| **R185W** | 160 | 186 | 70 | 87 | -3.516 | 0.025 |
| **R185W** | 163 | 177 | 6 | 36 | -3.618 | 0.022 |
| **R185W** | 164 | 299 | 8 | 25 | -5.792 | 0.004 |
| **R185W** | 169 | 377 | 46 | 9 | 4.046 | 0.016 |
| **R185W** | 170 | 377 | 88 | 33 | 7.927 | 0.001 |
| **R185W** | 172 | 177 | 95 | 70 | 3.896 | 0.018 |
| **R185W** | 183 | 263 | 24 | 13 | 5.821 | 0.004 |
| **R185W** | 340 | 345 | 53 | 39 | 4.306 | 0.013 |
| **R185W** | 344 | 349 | 43 | 29 | 5.604 | 0.005 |
| **R185W** | 348 | 376 | 38 | 0 | 3.312 | 0.03 |
| **R185W** | 348 | 377 | 87 | 20 | 11.32 | 0 |
| **R185W** | 349 | 354 | 98 | 65 | 2.936 | 0.043 |
| **R185W** | 349 | 355 | 91 | 60 | 4.729 | 0.009 |
| **R185W** | 352 | 356 | 89 | 68 | 5.686 | 0.005 |
| **R185W** | 353 | 357 | 94 | 51 | 4.696 | 0.009 |
| **R185W** | 354 | 376 | 57 | 1 | 3.234 | 0.032 |
| **R185W** | 354 | 377 | 27 | 10 | 5.099 | 0.007 |
| **R185W** | 356 | 375 | 0 | 53 | -3.947 | 0.017 |
| **R185W** | 357 | 374 | 59 | 3 | 4.13 | 0.014 |
| **R185W** | 357 | 376 | 93 | 26 | 15.169 | 0 |
| **R185W** | 358 | 374 | 59 | 4 | 4.426 | 0.011 |
| **R185W** | 358 | 375 | 0 | 54 | -3.857 | 0.018 |
| **R185W** | 359 | 374 | 70 | 8 | 5.349 | 0.006 |
| **R185W** | 359 | 375 | 33 | 87 | -3.402 | 0.027 |
| **R185W** | 359 | 376 | 1 | 53 | -6.749 | 0.003 |
| **R185W** | 360 | 374 | 16 | 2 | 3.156 | 0.034 |
| **R185W** | 360 | 375 | 0 | 16 | -3.055 | 0.038 |
| **R185W** | 363 | 374 | 79 | 30 | 5.195 | 0.007 |
| **R185W** | 363 | 375 | 0 | 48 | -10.399 | 0 |
| **R185W** | 367 | 374 | 18 | 52 | -3.945 | 0.017 |
| **R185W** | 370 | 374 | 42 | 95 | -4.137 | 0.014 |
| **R185W** | 371 | 375 | 35 | 91 | -3.582 | 0.023 |
| **R185W** | 372 | 376 | 0 | 67 | -15.053 | 0 |
| **R185W** | 373 | 376 | 0 | 67 | -12.103 | 0 |
| **R185W** | 373 | 377 | 0 | 26 | -4.84 | 0.008 |
| **G199S** | 25 | 347 | 51 | 40 | 3.158 | 0.034 |
| **G199S** | 41 | 51 | 13 | 1 | 4.313 | 0.013 |
| **G199S** | 44 | 48 | 25 | 72 | -4.363 | 0.012 |
| **G199S** | 45 | 48 | 21 | 74 | -3.325 | 0.029 |
| **G199S** | 45 | 49 | 24 | 80 | -3.674 | 0.021 |
| **G199S** | 46 | 51 | 24 | 6 | 6.68 | 0.003 |
| **G199S** | 104 | 131 | 38 | 28 | 3.412 | 0.027 |
| **G199S** | 200 | 248 | 17 | 41 | -4.916 | 0.008 |
| **G199S** | 259 | 263 | 69 | 81 | -3.538 | 0.024 |
| **R374S** | 3 | 6 | 20 | 65 | -3.318 | 0.029 |
| **R374S** | 3 | 30 | 39 | 7 | 3.421 | 0.027 |
| **R374S** | 4 | 355 | 2 | 15 | -4.836 | 0.008 |
| **R374S** | 7 | 349 | 32 | 18 | 3.239 | 0.032 |
| **R374S** | 7 | 355 | 64 | 41 | 2.83 | 0.047 |
| **R374S** | 8 | 24 | 30 | 65 | -5.097 | 0.007 |
| **R374S** | 8 | 104 | 89 | 78 | 3.383 | 0.028 |
| **R374S** | 39 | 53 | 85 | 50 | 2.845 | 0.047 |
| **R374S** | 41 | 65 | 74 | 87 | -3.406 | 0.027 |
| **R374S** | 43 | 51 | 47 | 7 | 3.417 | 0.027 |
| **R374S** | 44 | 51 | 27 | 2 | 3.829 | 0.019 |
| **R374S** | 45 | 53 | 0 | 14 | -4.01 | 0.016 |
| **R374S** | 48 | 51 | 39 | 9 | 4.221 | 0.013 |
| **R374S** | 51 | 54 | 13 | 58 | -5.033 | 0.007 |
| **R374S** | 51 | 82 | 8 | 51 | -3.546 | 0.024 |
| **R374S** | 51 | 85 | 1 | 29 | -9.405 | 0.001 |
| **R374S** | 51 | 86 | 6 | 51 | -5.115 | 0.007 |
| **R374S** | 52 | 86 | 22 | 63 | -2.852 | 0.046 |
| **R374S** | 56 | 83 | 35 | 13 | 2.895 | 0.044 |
| **R374S** | 100 | 104 | 39 | 24 | 3.798 | 0.019 |
| **R374S** | 100 | 125 | 19 | 7 | 4.532 | 0.011 |
| **R374S** | 104 | 131 | 38 | 23 | 3.052 | 0.038 |
| **R374S** | 106 | 348 | 49 | 75 | -3.559 | 0.024 |
| **R374S** | 122 | 136 | 52 | 22 | 3.484 | 0.025 |
| **R374S** | 130 | 361 | 66 | 79 | -4.319 | 0.012 |
| **R374S** | 135 | 372 | 56 | 0 | 2.985 | 0.041 |
| **R374S** | 135 | 374 | 61 | 0 | 3.567 | 0.023 |
| **R374S** | 135 | 377 | 44 | 93 | -2.788 | 0.049 |
| **R374S** | 137 | 348 | 22 | 39 | -4.015 | 0.016 |
| **R374S** | 137 | 376 | 31 | 0 | 3.302 | 0.03 |
| **R374S** | 141 | 376 | 18 | 0 | 3.437 | 0.026 |
| **R374S** | 145 | 170 | 11 | 23 | -3.157 | 0.034 |
| **R374S** | 145 | 377 | 72 | 36 | 2.936 | 0.043 |
| **R374S** | 150 | 377 | 27 | 3 | 3.343 | 0.029 |
| **R374S** | 169 | 377 | 46 | 15 | 3.478 | 0.025 |
| **R374S** | 348 | 376 | 38 | 0 | 3.312 | 0.03 |
| **R374S** | 354 | 376 | 57 | 7 | 2.86 | 0.046 |
| **R374S** | 357 | 374 | 59 | 0 | 4.338 | 0.012 |
| **R374S** | 358 | 363 | 65 | 24 | 3.391 | 0.028 |
| **R374S** | 358 | 374 | 59 | 0 | 4.751 | 0.009 |
| **R374S** | 359 | 363 | 76 | 44 | 3.305 | 0.03 |
| **R374S** | 359 | 364 | 64 | 28 | 2.914 | 0.043 |
| **R374S** | 359 | 375 | 33 | 91 | -3.972 | 0.017 |
| **R374S** | 360 | 374 | 16 | 0 | 3.604 | 0.023 |
| **R374S** | 363 | 374 | 79 | 2 | 11.273 | 0 |
| **R374S** | 370 | 374 | 42 | 91 | -3.838 | 0.018 |
| **R374S** | 371 | 375 | 35 | 96 | -4.043 | 0.016 |
| **R374S** | 372 | 375 | 43 | 100 | -2.856 | 0.046 |
